# Nutrition scores and MUAC of adult surgical orthopaedic inpatients at a teaching hospital in Lusaka province, Zambia

**DOI:** 10.1101/2022.08.10.22278631

**Authors:** Nixon Miyoba, Irene Ogada

## Abstract

**Background:** Poor nutrition status among hospitalised patients has been shown to increase length of hospital stay, as well as contribute to increased morbidity and mortality. The purpose of the study was to evaluate the nutrition status of adult surgical orthopaedic patients attending a teaching hospital in Zambia.

**Methods:** This study adopted a hospital-based cross-sectional study design to collect data from 98 adult patients aged 18 - 64 years. A structured questionnaire, the Subjective Global Assessment (SGA) tool, mid-upper arm circumference (MUAC) tape were used to collect data during the study period of three months in 2015.

**Results:** The mean age of the patients was 36.4 plus or minus 9.44 years, while the mean length of hospital stay was 17.33 plus or minus 10.91 days. Nutrition-focused physical examination revealed that majority (89.8%) of the patients were of acceptable weight with no weight loss reported in 70.4% of the patients. Poor appetite was only reported by 10.2% of the patients. SGA findings suggest that most of the patients (79.6%) were well-nourished. The mean mid-upper arm circumference of the study participants during hospitalization was 25.09 plus or minus 2.85 cm. An association was found between length of hospital stay and mid-upper arm circumference of the patients (p<0.001).

**Conclusion:** Subjective Global Assessment has the potential to evaluate the nutrition status of surgical patients in resource-poor settings such as Zambia. However, the use of SGA should be supplemented by other tools such as MUAC which has the potential to screen for adult malnutrition in clinical settings with limited resources.

## Background

Studies have shown that malnutrition in hospitals is a huge problem globally among surgical patients and is common among orthopaedics [1, 2, 3]. Malnutrition refers to a situation of deficiency, excess or imbalance of energy, protein and other nutrients resulting in adverse effects on functionality, body tissue and clinical outcomes [4]. In developed and developing countries, hospital malnutrition is prevalent [5]. For example, the prevalence of hospital malnutrition in most of the European countries is estimated at 37% [6]. A recent study conducted in regional hospitals in Ethiopia, Africa, revealed a prevalence of 55.6% [7].

In a study conducted in Paraguay, it was reported that malnutrition is a common condition on admission among trauma patients. According to the study, trauma was the leading cause of hospital admission of young people [8]. A study in a tertiary hospital in Spain on nutritional control revealed that the nutrition status of patients with hip and knee prosthesis deteriorated while in hospital [9]. It has been noted that the risk of malnutrition among patients increases while in hospital [3]. Therefore, a patient who presents with normal nutrition status may develop malnutrition if they stay longer in hospital. Malnutrition can thus lead to longer hospital stay [6, 10], and at the same time, longer hospital stay can result in malnutrition.

Among orthopaedic patients, malnutrition can impact bone health resulting in delayed healing [2]. During hospitalization some patients develop malnutrition while in hospital [11]. Moreover, identification of patients at risk of malnutrition is important to help in the provision of adequate nutritional therapy [12]. Hospital meals constitute an important component of patients’ care and a good nutrition status leads to faster recovery from illness [13]. Poor nutrition status on the other hand when associated with severity of disease condition has the potential to increase length of hospital stay and contribute to morbidity [14]. Adverse clinical outcomes may result if poor nutrition status is associated with delayed and insufficient post-operative nutrition [15].

Despite malnutrition being acknowledged to be common in hospitals, the condition is often ignored [16]. Literature points out that hospital malnutrition is frequently not detected and that recognition of malnutrition among inpatients still remains a challenge [17]. There is limited literature in Zambia and specifically Lusaka province on the nutrition status of adult surgical orthopaedic inpatients. Hence the purpose of the study was to evaluate the nutrition status of the orthopaedic patients.

## Methods

### Research design, period and location

The study adopted a cross-sectional study design with quantitative approaches to data collection, analysis and presentation. The design was appropriate as data was collected at one point in time [18]. The study was conducted from June to October 2015 at a teaching hospital in Lusaka province, Zambia. The teaching hospital is the main trauma referral centre in the country and is the largest public hospital, with a bed capacity of 1,655 [19, 20].

### Study population

The inclusion criteria for participating in the study were: adult surgical orthopaedic patients who were 18 – 64 years, admitted in low cost wards and able to verbally communicate [18, 21]. The study excluded patients who declined to participate, those who were cognitively impaired, as well as those who were critically ill [18]. Low cost wards are affordable for patients of low socio-economic status because it has cheaper medical services. Orthopaedic patients constituted 20 – 40% of the total admissions in surgical wards at the health facility [22].

### Sampling techniques and sample size

The teaching hospital was purposively sampled as it is the main trauma referral centre in the country [19, 20]. Participants were purposively drawn from the low cost surgical wards because that is where majority are admitted. After the nurses-in charge generated the sampling frame based on the admission records, comprehensive sampling was used to include all participants who met the inclusion criteria and consented to participate. A total of 98 participants were recruited into the study.

### Research instrument

It is recommended that malnutrition should be identified before it can be treated [23]. Several nutrition screening tools have been developed for use in the clinical setting but no tool is considered to be the yardstick for identifying malnutrition [24, 25, 26]. The nutrition screening tool must be simple, easy-to-use and patient-friendly [23]. A structured questionnaire with three components was used to collect data. The first section gathered demographic data (age, sex, marital status and days spent in hospital). The second part of the questionnaire elicited information using the Subjective Global Assessment (SGA).

SGA is a subjective tool used to categorise patients as well-nourished (Graded A), mildly/moderately malnourished (Graded B) and severely malnourished (Graded C). The basis for the classification is a focused history and physical examination [27, 28]. The components of the SGA are weight change, dietary intake change, gastrointestinal symptoms, functional capacity, disease effect and physical examination [27, 28].

Nutrition-focused physical assessment (NFPA) has been used to categorise the nutrition status of patients [29]. This is useful in situations where Body Mass Index (BMI) cannot be taken and weight loss categories are not possible [29]. A standard non-elastic mid-upper arm circumference (MUAC) tape was used to measure the MUAC of the participants during hospitalization. Results of MUAC were recorded in the last part of the research instrument. Prior to data collection, the instruments were pre-tested among adult surgical orthopaedic patients with similar characteristics but who were not included in the study.

### Data collection procedures

Prior to data collection, investigators were oriented to all low cost surgical wards by the senior nutritionist at the hospital. The nurse-in-charge in each ward assisted in obtaining informed consent from each patient and also made available the admissions books and patients’ files. Data collection started on a randomly selected day. A Medical Doctor in consultation with a Registered Dietitian collected data related to aspects of the Subjective Global Assessment (SGA).

Mid-upper arm circumference was measured on a randomly selected day. Anthropometric measurements of mid-upper arm circumference were taken by the investigators using a standard non-elastic MUAC tape to the nearest 0.1 cm. The measurement was taken half way between the acromion process of the scapula and the olecranon process at the tip of the elbow. Measurements were taken thrice while patients were seated on hospital beds. Any measurements that differed by more than 0.02 cm were discarded and new measurements were taken. The average of the three measurements was computed and recorded. MUAC has been used to assess adult weight change and to determine the prevalence of adult under-nutrition [29].

### Statistical analysis

All completed questionnaires were checked for accuracy, edited, cleaned and coded on a daily basis. Statistical Package for Social Sciences (SPSS) version 21.0 was used for data entry and analysis. Descriptive statistics in terms of means, frequencies, percentages and standard deviations were generated. Inferential statistics such as Pearson product moment coefficient were employed to establish relationships and a p-value of less than 0.05 was considered to be significant.

## Results

Out of the 104 adult surgical orthopaedic patients eligible to participate in the study, only 4 declined to participate. Analysis was based on data collected from 98 patients with completed data.

### Demographic characteristic of the patients

The mean age of the study participants was 36.4±9.44 years, while the mean length of stay in hospital was 17.33±10.91, ranging from 5 to 60 days (Table 1).

**Table 1:**
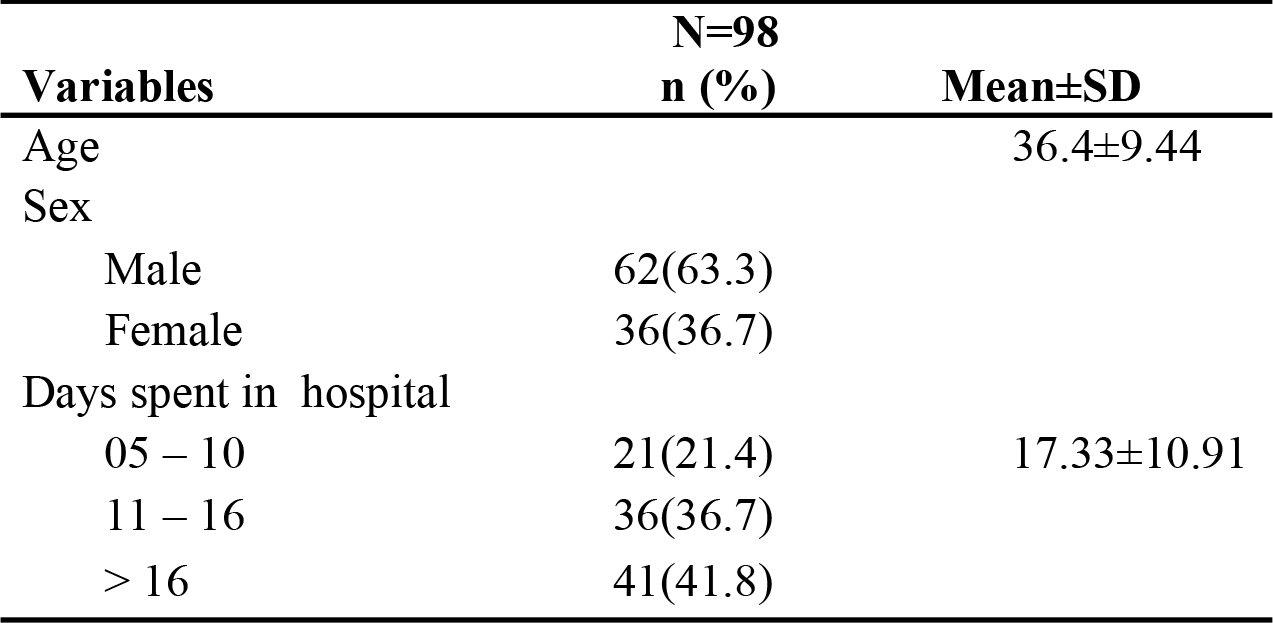
Demographic characteristic of the study participants.

### Appetite among the patients

Results of this study (Table 2) suggest that while the majority of the patients (89.8%) reported to have good and normal appetite, slightly less than 30% had poor appetite.

**Table 2:** Appetite of the study participants Appetite n (%)

| Appetite | n (%) |
| --- | --- |
| Normal | 60 (61.2) |
| Good | 28 (28.6) |
| Poor | 10 (10.2) |

### Body weight changes of the patients during hospitalization

Patients were asked about weight change during hospitalization. As depicted in Figure 1, majority of them (70.4%) indicated that their body weight had not changed.

**Figure 1:**
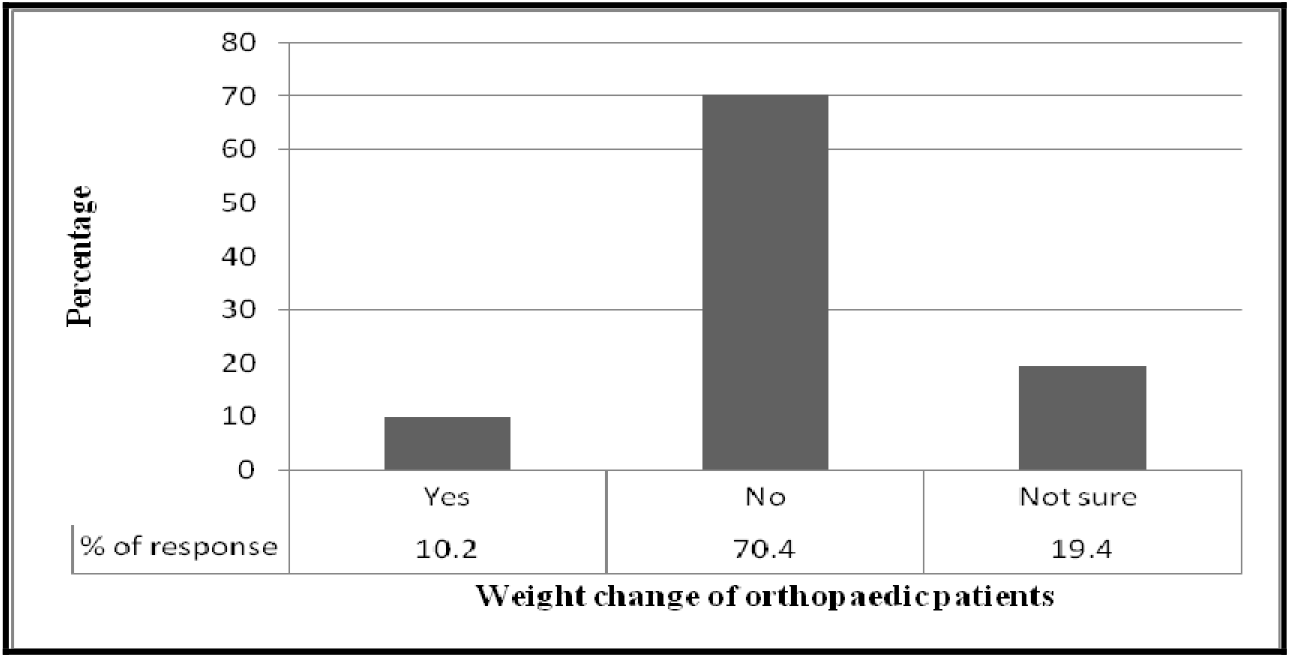
Patients’ reporting change in body weight during hospitalization.

### Nutrition-focused physical assessment (NFPA)

The nutrition-focused physical assessment revealed that most of the patients (89.8%) were of acceptable weight (Figure 2). Only 4.1% of the patients were either overweight or obese.

**Figure 2:**
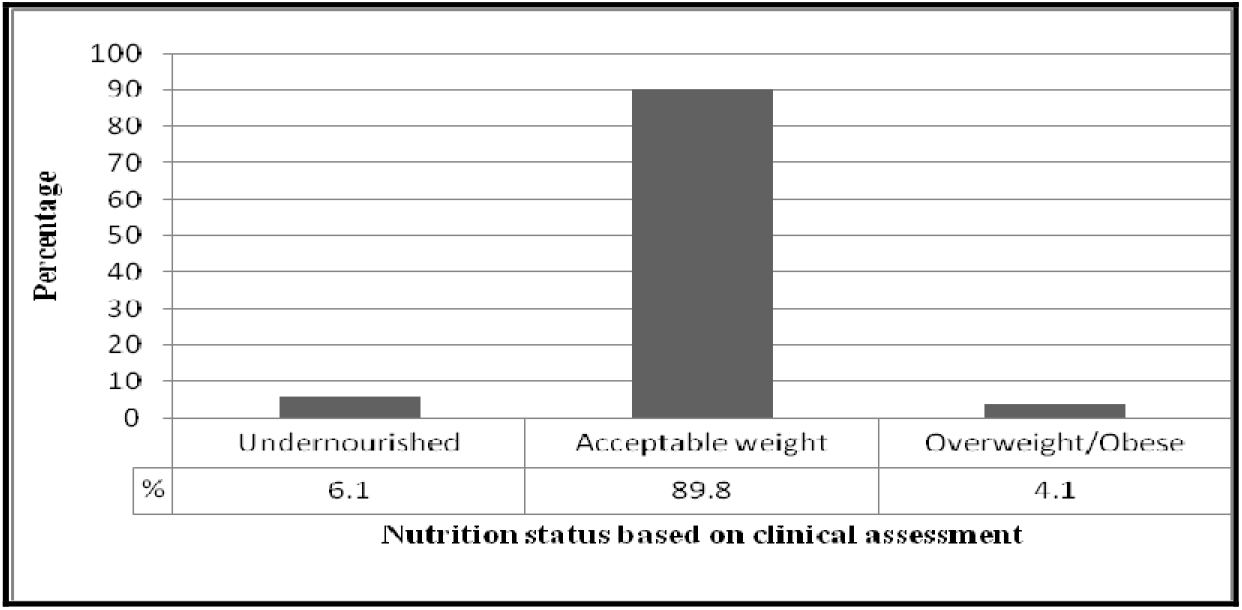
Nutrition status of the patients based on NFPA.

### Nutrition status of the participants as determined by MUAC

The majority (82.7%) of the patients were of normal nutrition status with 2.0% of them being either overweight or obese (Table 3). The mean mid-upper arm circumference of the patients was 25.09±2.85 cm. Classification was based on cut-offs used in other studies [29, 30, 31].

**Table 3:** MUAC measurements during hospitalization.

| MUAC | N=98<br>n(%) | Nutrition status | p-value |
| --- | --- | --- | --- |
| < 23 cm | 15(15.3) | Under nutrition |  |
| 23 ≤ to 32 cm | 81(82.7) | Normal | 0.001 |
| > 32 cm | 2(2.0) | Overweight/Obese |  |

A significant relationship was found between length of stay in hospital and MUAC of the patients (Pearson product moment correlation, $r=0.324$ , $p<0.001$ ).

### Subjective Global Assessment (SGA) scores of the patients

Results of the Subjective Global Assessment (SGA) indicate that 79.6% of the patients were well-nourished while 20.4% of them were either mildly or severely malnourished (Table 4).

**Table 4:** Classification of nutrition status of the patients.

| SGA categories | Grades | N=98<br>n(%) |
| --- | --- | --- |
| Well nourished | (A) | 78(79.6) |
| Mildly malnourished | (B) | 13(13.3) |
| Severely malnourished | (C) | 7(7.1) |

## Discussion

Evidence suggests that malnutrition among surgical patients greatly contributing to morbidity and mortality [1]. Adverse clinical outcomes may result if poor nutrition status is associated with insufficient pre- and post-operative nutrition [15]. Malnutrition can result in anaemia, hypoalbuminemia, impaired immunity and increased complications [32, 33, 34]. However, provision of adequate nutrition through the use of enteral and/or parenteral feeds has been shown to lower complications and mortality among patients with poor nutrition status [1, 35].

Most of the study participants (79.6%) were well-nourished. The SGA findings were supported by MUAC measurements which also suggested that most patients had normal nutrition status. Findings of this study are in congruence with those of a study in the United States of America [36]. The study revealed that the overall incidence of malnutrition among orthopaedic patients was low [36]. Similar to our findings, results of a study among patients undergoing Gastro-instestinal surgery suggested that 69% of the patients were well-nourished [27]. However, a study conducted in Paraguay among trauma patients reported that malnutrition is a common condition [8]. The variation in the incidence of under-nutrition may be attributed to the study site, patient group and assessment tool used to evaluate nutrition status.

The SGA first described in 1982 is applicable in clinical settings to evaluate the nutrition status of patients at the bedside [28]. This study collected data from patients who were not ambulant and hence the use of the SGA was appropriate. Owing to the fact that BMI cannot be computed for most of the orthopaedic inpatients, there is a high risk of them being ignored by nutrition practitioners in low-income countries. Yet, the SGA has been used widely to perform nutrition assessment among surgical inpatients [28]. The tool has been described to be simple to use, has the ability to assess functional status and has a high predictive value for mortality [28]. Literature suggests that even without anthropometric data, the SGA is still regarded a superior nutritional indicator [37].

In situations where Body Mass Index and weight loss categories cannot be taken, alternative anthropometric indicators such as MUAC can be used [29]. MUAC is easier to use, quicker, inexpensive and a useful nutritional indicator in both children and adults [38, 39, 40]. In low-income countries, MUAC is frequently used in community settings for rapid screening and surveillance programs [38]. For example, in a study conducted in Sudan, nutrition assessment was done with MUAC and its results and correlated well with BMI of adults [41].

The current study found a correlation between length of stay and MUAC of patients. Findings indicated a significant association between the two variables (P<0.001). A study on the feasibility of MUAC as a screening tool in tertiary public hospitals in South Africa only reported on the significant relationship between MUAC, BMI and nutrition scores [40]. There is limited literature in hospitals in the sub-Saharan Africa on associations between MUAC and length of hospital stay. However, the use of MUAC has been reported in resource-poor settings and is useful in clinical settings to evaluate the nutrition status of patients [40, 42].

We recommend the use of the Subjective Global Assessment (SGA) tool to evaluate the nutrition status of surgical patients in resource-poor settings. This can be done by a Medical Doctor working in collaboration with nutrition professionals. Identification of malnutrition among surgical patients may contribute to early formulation and implementation of appropriate nutrition therapies. Results of the SGA can be supplemented with MUAC findings to aid nutrition professionals develop sound clinical judgment. A multi-center study is recommended to assess the feasibility of SGA among surgical patients in Zambia and correlate its findings with clinical outcomes.

## Study limitations

Results of the study cannot be generalized to all public hospitals in the country as data was only collected from one group of patients at a university teaching hospital. The other limitation was the use of only one anthropometric tool to evaluate the nutrition status of the patients. However, this shortcoming was overcome by use of the Subjective Global Assessment (SGA) tool. Despite the limitation in data, this study has provided some useful information that may assist nutrition practitioners in developing countries to devise ways to evaluate the nutrition status of surgical patients.

## Conclusion

MUAC is associated with length of hospital stay. Subjective Global Assessment has the potential to evaluate the nutrition status of surgical patients in resource-poor settings such as Zambia. However, the use of SGA should be supplemented by other tools such as MUAC which has the potential to screen for adult malnutrition in clinical settings with limited resources.

## Data Availability

Data is available upon request from the corresponding author in consultation with the research institution.

## Ethical considerations

Ethical approval was obtained from Eres Converge (Number.2015-May-014) and permission to conduct the study was obtained from the University Teaching Hospital management. The purpose and processes of the study, as well as participants’ rights were clearly explained to the study participants prior to data collection. Participation in the study was voluntary and informed consent was obtained from the study participants prior to participation.

## Acknowledgement

The authors would like to thank management and staff at the hospital for the support rendered during the course of the study. In addition, the investigators express their gratitude to Mwape Chanda and Pharoah Banda for their key role during data collection.

## References

[1] Ben-Ishay, O., Gertsenzon, H., Mashiach, T., Kluger, Y., & Chermesh I. (2011). Malnutrition in surgical wards: A plea for concern. Gastroenterology Research and Practice, 1 – 4.

[2] Deren, M. E., Huleatt, J., Winkler, M. F., Runbin, L. E., & Salzler, M. J. (2014). Assessment and treatment of malnutrition in orthopaedic surgery. Journal of Bone and Joint surgery, 1 – 10.

[3] Gottraux, S., Maisonneuve, M., Gevaux, D., Fonzo-Christe, C., Chiki, M., Guinot-Bourquin, S., et al. (2004). Screening and treatment of malnutrition: European Council Resolution and its potential application in Switzerland. Revue medicale de la Suisse romande, 124(10), 617–623.

[4] Elia, M. (2003). Screening for malnutrition: a multidisciplinary responsibility. Development and use of the ‘Malnutrition Universal Screening Tool’ (‘MUST’) for adults. Redditch: BAPEN.

[5] Kim, K., Kim, M., & Lee, K. (2010). Assessment of food servive quality and identification of improvement strategies using hospital food service quality model. Nutr Res Pract, 4(2), 163 – 172.

[6] Kondrup, J., & Sorensen, J. (2009). The magnitude of the problem of malnutrition in Europe. Copenhagen: Nestec Ltd.

[7] Haile, A., Hailu, M., & Tesfaye, E. (2015). Prevalence and associated factors of malnutrition among adult hospitalized patients at Amhara National Regional State Referral Hospitals, Ethiopia. Integr Obesity Diabetes, 1(3), 80 – 83.

[8] Goiburu, M. E., Goiburu, M. M., Bianco, H., Ruiz Diaz, J., Alderete, F., Palacios, M. C., et al. (2006). The impact of malnutrition on morbidity, mortality and length of hospital stay in trauma patients. Nutr Hosp, 6004 – 610.

[9] Garcia, D. S., Perez, S. G., Sanavia, M. E., de Juanes, P. J., Arrazola, M. M., & Resines, E. C. (2008). Nutritional control in orthopaedic surgery patients. Nutr Hosp, 493 – 499.

[10] Thompson, B. A. (2013). Hand grip strength (HGS) as an indicator of nutritional status in patients in a rural hospital. Bega: Rural Research Capacity Building Program 2011 intake.

[11] Kirkland, L.L., Kashiwagi, D.T., Brantley, S., Scheurer, D., Varkey, P. (2013). Nutrition in the hospitalized patient. J Hosp Med. 8(1), 52 – 58

[12] Aquino, R., & Philipi, S. (2011). Identification of maluntrition risk factors in hospitalized patients. Rev Assoc Med Bras, 623 – 629.

[13] Mentziou, I., Delezos, C., Nestoridou, A., & Boskou, G. (2014). Evaluation of food services by the patients in hospital of Athens in Greece. Health Science Journal, 383–392.

[14] ESPEN. (2003). ESPEN guidelines for nutrition screening. Clin Nutri, 415–421.

[15] Garth, A. K., Newsome, C. M., Simmance, N., & Crowe, T. C. (2010). Nutritional status, nutrition practices and post-operative complications in patients with gastrointestinal cancer. Journal of Human Nutrition and Dietetics, 393 – 401.

[16] Tappenden, K. A., Quatrara, B., Parkhurst, M. L., Malone, A. M., Fanjiang, G., & Ziegler, T. R. (2013). Critical role of nutrition in improving quality of care: An interdisciplinary call to action to address hospital malnutrition. J Acad Nutr Diet, 1219 –1237.

[17] Norman, K., Pichard, C., Lochs, H., & Pirlich, M. (2008). Prognostic impact of disease - related malnutrition. Clinical Nutrition, 5 – 15.

[18] Miyoba, N., Ogada, A. I. & Mulenga, J., (2018). Dietary adequacy of adult surgical orthopaedic patients admitted to a teaching hospital in Lusaka province Zambia; A hospital-based cross-sectional study. BMC Nutrition. 4(37), 1 – 8.

[19] Kinnear J, Bould MD, Ismailova F., & Measures E. (2013). Can A new partnership for anesthesia training in Zambia: Reflections on the first year. J Anesth. 60(5), 1–8.

[20] Zambia UK Health Workforce Alliance. (2013). Retrieved from https://www.ed.ac.uk/global-health/research-projects-communities-of-practice/themes/healthequity-and-equality/zukhwa.

[21] McPake, B., Nakamba, P., Hanson, K., & McLoughlin, B. (2004). Private wards in public hospitals: two-tier charging and the allocation of resources in tertiary hospitals in Zambia. Lusaka, Zambia.

[22] University Teaching Hospital. (2015). Admission records. Lusaka: University Teaching Hopsital.

[23] Velasco, C., Garcia, E., Rodriguez, V., Frias, L., Garriga, R., Alvarez, J., et al. (2011). Comparison of four nutritional screening tools to detect nutritional risk in hospitalized patients: a multicenter study. European Journal of Clinical Nutrition, 269 – 274.

[24] Cant, R. P. (2011). Investing in patients’ nutrition: Nutrition risk screening in hospital. Australian Journal of Advanced Nursing, 81 – 87.

[25] Lomivorotov, V. V., Efremov, S. M., Boboshko, V. A., Nikolaev, D. A., Vedernikov, P. E., & Karaskov, A. M. (2013). Evaluation of nutritional screening tools for patients scheduled for cardiac surgery. Nutrition, 436 – 442.

[26] Sakinah, H., Suzana, S., Noor, A. M., Philip, P. J., & Shahrul, B. K. (2012). Development of a Local Malnutrition Risk Screening Tool-Hospital (MRST-H) for hospitalized elderly patients. Mal J Nutr, 137 – 147.

[27] Detsky AS, Baker JP, O’Rourke K, et al. (1987). Predicting nutritionassociated complications for patients undergoing gastrointestinal surgery. JPEN J Parenter Enteral Nutr. 11, 440 – 446.

[28] Makhija, S., & Baker, J. (2008). The Subjective Global Assessment: A Review of Its Use in Clinical Practice. Nutr Clin Pract. 4, 405 – 409.

[29] The Parenteral and Enteral Nutrition Group. (2011). A pocket guide to clinical nutrition. London : The Parenteral and Enteral Nutrition Group of the British Dietetic Association.

[30] Bisai, S. (2009). Undernutrition in the Kora Mudi Tribal Population, West Bengal, India: a comparison of body mass index and mid-upper-arm circumference. Food & Nutrition Bulletin, 30(1), 63–7.

[31] Chakraborty, R. (2009). Mid-upper arm circumference as a measure of nutritional status among adult bengalee male slum dwellers of Kolkata, India: relationship with self-reported morbidity. Anthropologischer Anzeiger, 67(2), 129–37.

[32] Harris, D. G., Davies, C., Ward, H., & Haboubi, N.Y. (2008). “An observational study of screening for malnutrition in elderly people living in sheltered accommodation,” Journal of Human Nutrition and Dietetics, 21(1), 3 – 9.

[33] Osada, J., Kamocki, Z., Rusak, M., Dabrowska, M., & Kedra, B. (2008). “The effect of surgical and nutritional treatment on activation parameters of peripheral blood T lymphocytes in stomach cancer patients in post operative period. Polski Merkuriusz Lekarski, 24(141)231236.

[34] Collins, C.E., Kershaw, J., & Brockington, S. (2005). “Effect of nutritional supplements on wound healing in home-nursed elderly: a randomized trial,” Nutrition, 21(2)147 – 155.

[35] ASPEN (2002). Board of Directors and the Clinical Guidelines Task Force, “Guidelines for the use of parenteral and enteral nutrition in adult and pediatric patients,” Journal of Parenteral and Enteral Nutrition, 26(1)1SA–138SA.

[36] Huang, R., Greenky, M., Kerr, G., Austin, M., & Parvizi, J. (2013). The effect of malnutrition on patients undergoing elective joint arthroplasty. The Journal of Arthroplasty, 21 – 24.

[37] Planas, M., Audivert, S., Perez-Portabella C, et al. (2004). Nutritional status among adult patients admitted to a university-affiliated hospital in Spain at the time of genoma. Clin Nutr. 23, 1016 – 1024.

[38] Roy, N.C. (2000). Use of mid-upper arm circumference for evaluation of nutritional status of children and for identification of high-risk groups for malnutrition in rural Bangladesh. J Health Popul Nutr. 18, 171 – 80.

[39] Tsai, A. C., Chang, T. L, Yang, T. W, Chang-Lee, S.N, Tsay, S.F. (2010). A modified mini nutritional assessment without BMI predicts nutritional status of community-living elderly in Taiwan. J Nutr Health Aging. 14, 183–9.

[40] Velzeboer, MI, Selwyn BJ, Sargent F, Pollitt E, Delgado H. (1983). The use of arm circumference in simplified screening for acute malnutrition by minimally trained health workers. J Trop Pediatr. 29, 159 – 66.

[41] Collins, S. (1996). Using middle upper arm circumference to assess adult malnutrition during famine. JAMA. 276(5): 391 – 395. PMID:8683818.

[42] Miyoba, N., Musowoya, J., Mwanza, E., Malama, A., Murambiwa, N., Ogada I., Njobvu M & Liswaniso D. (2018). Nutritional risk and associated factors of adult in-patients at a teaching hospital in the Copperbelt province in Zambia; a hospital-based cross-sectional study. BMC Nutrition. 4(40), 1 – 6.

